# Determinants of wasting, stunting, and undernutrition among children under five years: Cross-sectional study in southern Punjab, Pakistan

**DOI:** 10.1101/2023.01.04.23284177

**Authors:** Javeria Saleem, Rubeena Zakar, Rana Muhammad Aadil, Muhammad Salman Butt, Faisal Mushtaq, Gul Mehar Javaid Bukhari, Florian Fischer

## Abstract

**Background:** Malnutrition is a serious concern globally and may lead to early death if it remains untreated. Prevalence of malnutrition is high in South Asian countries. Therefore, this study aims to evaluate the determinants of wasting, stunting, and undernutrition in under-five children of southern Punjab, Pakistan.

**Methods:** We conducted a cross-sectional study among 185 children. Anthropometric measures were done by nutritional experts and pediatricians. Data were analyzed with SPSS version 25.0. A p-value <0.05 was considered statistically significant.

**Results:** Significant determinants of wasting (weight-for-height) were family monthly earnings (β=-0.14; 95% CI: -0.89 to -0.04; p=0.03) and complementary feeding practices (β=-0.21; 95% CI: -1.14 to 0.19; p<0.001). For stunting (length/height-for-age), the significant determinants were tuberculosis (TB) contact history (β=-0.15; 95% CI: -0.97 to -0.03; p=0.03) and non-use of exclusive breastfeeding practices (β=-0.19; 95% CI: -1.40 to 0.16; p=0.01). For undernutrition, significant determinants were monthly income (β=0.28; 95% CI: 0.11 to 0.62; p=0.02) and exclusive breastfeeding practices (β=0.22; 95% CI: 0.17 to 0.39; p=0.02).

**Conclusion:** Social determinants such as family earnings, family food security, practices of exclusive breastfeeding and proper complementary feeding, number of under-five siblings, and history of TB contact have a strong association with malnutrition and undernutrition. Concerted and comprehensive strategies are needed for the improvement of associated factors to combat malnutrition as well as undernutrition among children.

## Introduction

Malnutrition with all its types, especially undernutrition, wasting (height-for-weight), and stunting (height-for-age) are usually characterized by abnormal or inadequate food intake (1). Owing to undernutrition, children are more susceptible to diseases or even death. The risk of death increases in moderate to severely wasted children. According to the World Health Organization (WHO), stunting is also present among children with low socioeconomic status, poor feeding practices, poor nutrition and health of the mother, and child illness in the early stages of life [1].

Globally, the situation of malnutrition is critical: In 2019, 47.0 million children under the age of five years were wasted and 144.0 million children were stunted. South Asia has the highest number of wasted (25.2 million; 14.3% of children under five in South Asia) and stunted (55.9 million; 31.7% of children under five in South Asia) children under five [2]. Overall, families with higher income and education levels had lower rates of underweight children [3].

The death rate associated with undernutrition is higher in low-to middle-income countries. Almost 45% of deaths of children below the age of five years were associated with undernutrition [1]. Wasted children are more prone to experience several infections, delays in cognitive development, and reduced productivity in adulthood [4]. The economic, medical, social as well as developmental impacts of malnutrition cause decreased growth and development potential of children. It is a very serious issue not only for individuals and families, but also for economic productivity of already resource scare countries [1].

According to the National Nutrition Survey 2018, Pakistan has a 40.2% stunting prevalence, 17.7% wasting rate, and 28.9% of children below the age of five years are underweight. Overall, 48.4% of infants received exclusive breastfeeding and 45.8% of mothers started early initiation of breastfeeding. Boys are slightly more stunted, wasted, and underweight (40.9%, 18.4%, and 29.3%, respectively) as compared to girls (39.4%, 17.0%, and 28.4%, respectively) [5]. Although a growing body of research on child nutritional status is available in Pakistan, studies are still lacking which investigate the effect of different determinants of malnutrition. Therefore, the purpose of the current study was to understand the determinants of wasting, stunting, and undernutrition to identify new targets for intervention to prevent malnutrition of children under five years of age in southern Punjab, Pakistan.

## Methods

### Study design

A multicenter cross-sectional study was conducted at Community Management of Acute Malnutrition (CMAM) centers in four villages of southern Punjab, running under the supervision of the health department of the government of Pakistan. These were government-designated centers for the management of malnutrition in under five children. Study participants enrolled from these centers visit CMAM centers to receive their nutritional treatment. Southern Punjab is a highly populated and socio-economically deprived area with the worst indicators of illiteracy, malnutrition, infant and under-five child mortality in Pakistan [5,6].

### Sample

The sample size was calculated by using the cross-sectional study formula taking severe malnutrition prevalence (p) as 11% and error term (d) as 0.05. The calculated sample size was n=151. This number was further enhanced to 185 for study precision, strength and accuracy. A total of 185 children aged 6–59 months without gender specification were enrolled. Eligibility criteria were having severe wasting or stunting identified by anthropometric indices of WHO classification, which are having height-for-weight (wasting) < -3 standard deviations (SD) and height-for-age (stunting) < -2 SD [7,8]. All children were from the same ethnic and language background. Written consent was obtained from parents after introducing them to the purpose of the study. A flow chart for eligibility criteria is presented in Figure 1.

**Figure 1:**
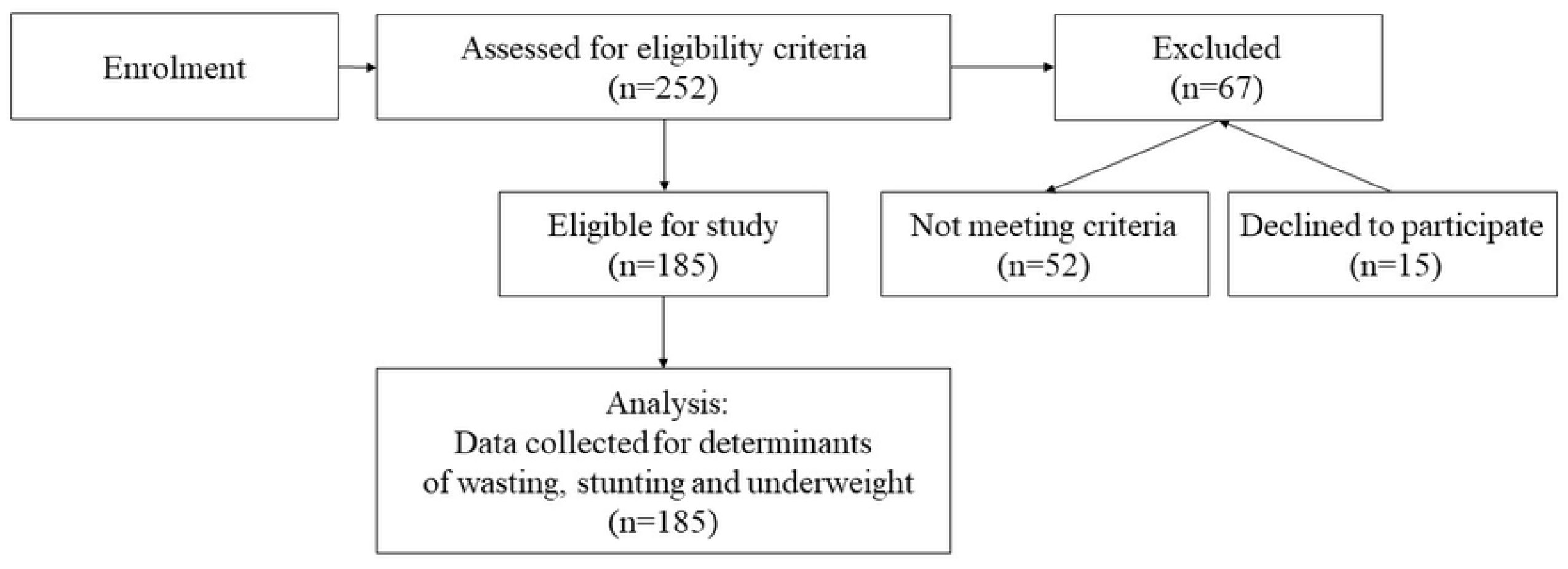
Flowchart of participant recruitment.

### Baseline assessments

Interviews with parents were conducted at CMAM centers. An extensively pretested structured questionnaire was used for collection of information from parents/caregivers regarding family sociodemographic characteristics (income, parental education, profession, household size, and family structure), child nutrition (breastfeeding and complementary feeding practices), biological and healthcare characteristics (child medical history, immunization record, and access to health care). Family monthly earning was assessed by income from all sources in Pakistani rupees (PKR). Exclusive breastfeeding and complementary feeding practices (quantity, variety, and frequency) of semisolid diet were judged according to WHO recommendations [9]. For immunization status, the Expanded Program of Immunization (EPI) schedule was followed [10]. The immunization status was verified by the vaccination card. Child medical history was assessed by previous hospital records and prescriptions. A brief history was acquired from parents/caregivers to identify the child for any seizure disorder and neurological deficit. Children sick with these conditions were not enrolled in the study. All of these determinants of wasting, stunting, and underweight were included in the study after an extensive literature search.

In case of hospital delivery, child gestational age was calculated from previous antenatal hospital documentation and in case of delivery at home, information based on a maternal report was noted. For children who were delivered before 37 weeks of gestation and at the date of data colletion were ≤24 months of age, their age was adjusted by deducting the entire weeks of missed gestation from the current age.

### Anthropometric assessments

Anthropometric assessments were taken by the qualified nutritional supervisors. Their competency in assessing weight and height was approved by the first author. Double anthropometric values were recorded by a study team. If both values were different from each other, then further measurements were conducted until an exact value was measured. The replicated value was then documented. Children’s weight (approximated to 10 g) was assessed without clothing or in very light outfit through a UNISCALE, which was fixed by a standard weight and adjusted to zero before each evaluation. First, UNISCALE was used to record only the weight of the mother if infants and children were not able to stand. Afterwards, the undressed baby or child was given to the mother as she stood on the scale, and the cumulative weight of the mother and infant was evaluated [8,11]. Infant/child weights were calculated as the difference between these two measures.

The recumbent length of kids ≤ 87 cm in height was measured to the nearest 0.1 cm with a length-measuring board with a fixed headrest and an adjustable foot piece (“SECA GmbH & Co. KG, Hamburg, Germany”). Children over 87 cm tall were evaluated by standing with their bare feet and heels together on a flat horizontal plate attached to the measurement board. Weight-for-height Z-scores and height-for-age Z-scores were calculated using the WHO Child Growth Standards [8,11]. Nutrition status classification was done with “WHO ANTHRO, version 3.2.2”.

### Dependent and independent variables

In this study analysis, the dependent variables were wasting (weight-for-height Z-score) as a measure of acute malnutrition, stunting (height-for-age Z-score) as a measure of chronic malnutrition, and undernutrition (weight-for-age Z-score). All sociodemographic, medical, and dietary factors described above have been included as independent variables.

### Data analysis

Data was analyzed using SPSS 25.0. An independent t-test was used to compare the Z-score mean differences between independent variables for wasting, stunting, and undernutrition. A multivariate analysis was performed to identify the significant determinants causing wasting, stunting, and undernutrition among children under five years. Based on continuous dependent variables, Linear regression models were applied for multivariate analyses. The results were reported as β coefficients with 95% confidence intervals (CI). p-values of 0.05 and less were considered as statistically significant.

### Ethical approval

Permission to conduct the study was obtained from the provincial and district health department of Punjab, Pakistan. This study was conducted according to the guidelines laid down in the Declaration of Helsinki and all procedures involving human subjectss were approved by the Ethical Review and Advanced Study Research Board of the University of Punjab, Pakistan (ref-9/2352-ACAD). Written informed consent from parents was obtained from all subjects.

## Results

A total of 185 children aged 15.36 months (SD±10.23) participated in this study. They had a mid-upper arm circumference of 10.19 cm (SD±0.96). The majority were females (n=104; 56.2%), had a family income of ≤15000 PKR (n=128; 69.2%) and at least two siblings (n=144; 77.8%). Overall 100 children (54.1%) had one to seven previous episodes of illness, 149 (80.5%) had no exclusive breastfeeding, and poor complementary feeding practices were seen in 165 children (89.2%) children as shown in Table 1.

**Table 1:** Characteristics of study participants.

| Variable | Category | n (%) |
| --- | --- | --- |
| Age<br>(15.36 ± 10.23) | ≤ 12 months | 112 (60.5) |
|  | 13-24 Months | 45 (24.3) |
|  | ≥ 25 Months | 28 (15.1) |
| Gender | Male | 81 (43.8) |
|  | Female | 104 (56.2) |
| Income per month | ≤ 15,000 PKR | 128 (69.2) |
|  | 15,000-35,000 PKR | 57 (30.8) |
| Under-five siblings | ≤ 2 | 144 (77.8) |
|  | ≥ 3 | 41 (22.2) |
| Immunization | Incomplete | 46 (24.9) |
|  | Complete | 139 (75.1) |
| History of parasite | Yes | 42 (22.7) |
|  | No | 143 (77.3) |
| History of TB | Yes | 81 (43.8) |
|  | No | 104 (56.2) |
| History of measles | Yes | 28 (15.1) |
|  | No | 157 (84.9) |
| Episodes of Illness | 1-7 | 100 (54.1) |
|  | 8-15 | 85 (45.9) |
| Exclusive breastfeeding | Yes | 36 (19.5) |
|  | No | 149 (80.5) |
| Complementary feeding practice | Poor | 165 (89.2) |
|  | Good | 20 (10.8) |

### Determinants of wasting (acute malnutrition: weight-for-height Z-score)

In the bivariate analysis, wasting was significantly associated with family monthly earning with a mean difference of 0.43 (95% CI: 0.01 to 0.85; p=0.04) and complementary feeding practices with a mean difference of 0.51 (95% CI: 0.05 to 0.96; p=0.03). Applying linear regression for the multivariate analysis, family monthly income (β=-0.14; 95% CI: -0.89 to -0.04; p=0.03) and complementary feeding practices (β=-0.21; 95% CI: -1.14 to 0.19; p<0.001) were again significant determinants (Table 2).

**Table 2:** Determinants of wasting.

| Independent variables |  | Mean (SD) | t-test / Bivariate analysis<sup>[a]</sup> |  | Multivariate analysis<sup>[b]</sup> |  |
| --- | --- | --- | --- | --- | --- | --- |
|  |  |  | Mean difference<br>(95% CI) | p | β (95% CI) | p |
| Gender | Male | -3.89 (1.41) | 0.03<br>(-0.37 to 0.43) | 0.88 | -0.02<br>(-0.47 to 0.33) | 0.73 |
|  | Female | -3.92 (1.32) | Reference |  | Reference |  |
| Age in months | ≤12 | -3.85 (1.43) | Reference | 0.29 | Reference | 0.12 |
|  | 13-24 | -4.17 (1.07) | -0.32<br>(-0.89 to 0.25) |  | -0.12<br>(-0.89 to 0.11) |  |
|  | ≥25 | -3.71 (1.46) | 0.14<br>(-0.54 to 0.81) |  | 0.008<br>(-0.55 to 0.62) |  |
| Monthly earning<br>(PKR) | <15000 | -4.04 (1.36) | Reference | 0.04* | Reference | 0.03* |
|  | 15000-35000 | -3.61 (1.32) | 0.43<br>(0.01 to 0.85) |  | -0.14<br>(-0.89 to 0.04) |  |
| Under five<br>siblings | ≤2 | -3.98 (1.39) | Reference | 0.14 | Reference | 0.12 |
|  | ≥3 | -3.64 (1.25) | 0.35<br>(-0.12 to 0.82) |  | -0.11<br>(-0.85 to 0.10) |  |
| History of<br>Parasites | Yes | -3.88 (1.47) | 0.03<br>(-0.44 to 0.50) | 0.89 | -0.03<br>(-0.59 to 0.35) | 0.61 |
|  | No | -3.91 (1.33) | Reference |  | Reference |  |
| TB contact<br>history | Yes | -4.09 (1.34) | -0.31<br>(-0.71 to 0.08) | 0.09* | 0.14<br>(-0.01 to 0.77) | 0.05* |
|  | No | -3.77 (1.36) | Reference |  | Reference |  |
| Measles history | Yes | -4.07 (1.12) | 0.19<br>(-0.74 to 0.36) | 0.50 | 0.06<br>(-0.33 to 0.79) | 0.42 |
|  | No | -3.88 (1.40) | Reference |  | Reference |  |
| Immunization<br>status | Done | -3.84 (1.31) | Reference | 0.23 | Reference | 0.32 |
|  | Incomplete | -4.12 (1.49) | -0.28<br>(-0.74 to 0.18) |  | 0.07<br>(-0.23 to 0.70) |  |
| Episodes of<br>illness | 1-7 | -3.85 (1.27) | Reference | 0.58 | Reference | 0.13 |
|  | 8-15 | -3.97 (1.46) | 0.12<br>(-0.52 to 0.27) |  | 0.11<br>(-0.09 to 0.71) |  |
| Complementary<br>feeding practice | Poor | -4.03 (1.34) | Reference | 0.03* | Reference | <0.001* |
|  | Good | -3.53 (1.39) | 0.51<br>(0.05 to 0.96) |  | -0.21<br>(-1.14 to -0.19) |  |
| Exclusive<br>breastfeeding | Yes | -3.82 (1.32) | 0.10<br>(-0.39 to 0.60) | 0.67 | -0.01<br>(-0.56 to 0.47) | 0.87 |
|  | No | -3.92 (1.37) | Reference |  | Reference |  |

<sup>[a]</sup> Independent t-test for bivariate analysis
<sup>[b]</sup> Multivariate analysis
\* p-value <0.05

### Determinants of stunting (chronic malnutrition: height-for-age Z-score)

In bivariate analysis of stunting, tuberculosis (TB) contact history and the practices of exclusive breastfeeding with a mean difference of 0.41 (95% CI: -0.53 to 0.86; p=0.04) and 0.75 (95% CI: 0.18 to 1.31; p=0.01), respectively, were statistically significant. The linear regression showed a significant association of stunting with TB contact history (β=-0.15; 95% CI: -0.97 to 0.03; p=0.03) and exclusive breastfeeding practices (β=-0.19; 95% CI: -1.40 to 0.16; p=0.01) (Table 3).

**Table 3:** Determinants of stunting.

| Independent variables |  | Mean (SD) | t-Test / Bivariate analysis<sup>[a]</sup> |  | Multivariate analysis<sup>[b]</sup> |  |
| --- | --- | --- | --- | --- | --- | --- |
|  |  |  | Mean difference (95% CI) | p | β (95% CI) | p |
| Gender | Male | -3.93 (1.66) | -0.20<br>(-0.66 to 0.27) | 0.40 | 0.08<br>(-0.20 to 0.74) | 0.27 |
|  | Female | -3.73 (1.50) | Reference |  | Reference |  |
| Age in months | ≤12 | -3.86 (1.55) | Reference | 0.80 | Reference | 0.76 |
|  | 13-24 | -3.84 (1.61) | 0.02<br>(-0.64 to 0.68) |  | -0.02<br>(-0.68 to 0.50) |  |
|  | ≥25 | -3.64 (1.63) | 0.22<br>(-0.57 to 1.01) |  | 0.03<br>(-0.52 to 0.86) |  |
| Monthly earning (PKR) | <15,000 | -3.77 (1.26) | Reference | 0.54 | Reference | 0.20 |
|  | 15,000-35,000 | -3.92 (1.61) | -0.15<br>(-0.65 to 0.34) |  | 0.09<br>(-0.18 to 0.84) |  |
| Under five siblings | ≤2 | -3.80 (1.55) | Reference | 0.74 | Reference | 0.96 |
|  | ≥3 | -3.89 (1.66) | -0.09<br>(-0.64 to 0.46) |  | 0.003<br>(-0.55 to 0.58) |  |
| TB contact history | Yes | -3.59 (1.50) | 0.41<br>(-0.53 to 0.86) | 0.04* | -0.15<br>(-0.967 to 0.03) | 0.03* |
|  | No | -3.99 (1.61) | Reference |  | Reference |  |
| History of parasites | Yes | -3.93 (1.66) | -0.14<br>(-0.69 to 0.40) | 0.61 | 0.04<br>(-0.37 to 0.74) | 0.52 |
|  | No | -3.78 (1.55) | Reference |  | Reference |  |
| History of measles | Yes | -4.08 (1.40) | -0.30<br>(-0.94 to 0.32) | 0.34 | 0.04<br>(-0.44 to 0.88) | 0.52 |
|  | No | -3.77 (1.60) | Reference |  | Reference |  |
| Complementary feeding practices | Good | -3.80 (1.65) | 0.02<br>(-0.51 to 0.55) | 0.93 | 0.03<br>(-0.42 to 0.68) | 0.63 |
|  | Poor | -3.82 (1.55) | Reference |  | Reference |  |
| Exclusive breastfeeding | Yes | -3.22 (1.69) | 0.75<br>(0.18 to 1.31) | 0.01* | -0.19<br>(-1.40 to 0.16) | 0.01* |
|  | No | -3.96 (1.51) | Reference |  | Reference |  |
| Episodes of illness | 1-7 | -3.86 (1.64) | Reference | 0.68 | Reference | 0.50 |
|  | 8-15 | -3.77 (1.49) | 0.09<br>(-0.36 to 0.55) |  | -0.05<br>(-0.63 to 0.31) |  |
| Immunization status | Done | -3.83 (1.52) | Reference | 0.83 | Reference | 0.96 |
|  | Incomplete | -3.77 (1.74) | 0.05<br>(-0.47 to 0.58) |  | 0.01<br>(-0.53 to 0.56) |  |

<sup>[a]</sup> Independent t-test for bivariate analysis
<sup>[b]</sup> Multivariate analysis
\* p-value <0.05

### Determinants of undernutrition (weight-for-age Z-score)

The results for undernutrition are shown in Table 4. In the bivariate analysis, monthly income with a mean difference of 0.32 (95% CI: 0.03 to 0.61; p=0.03) and the practices of exclusive breastfeeding with a mean difference of 0.39 (95% CI: 0.05 to 0.73; p=0.03) were significantly associated with undernutrition. Application of linear regression in the multivariate analysis showed a statistically significant association between undernutrition and monthly income (β=0.28; 95% CI: 0.11 to 0.62; p=0.02) and exclusive breastfeeding practices (β=0.22; 95% CI: 0.17 to 0.39; p=0.02).

**Table 4:**
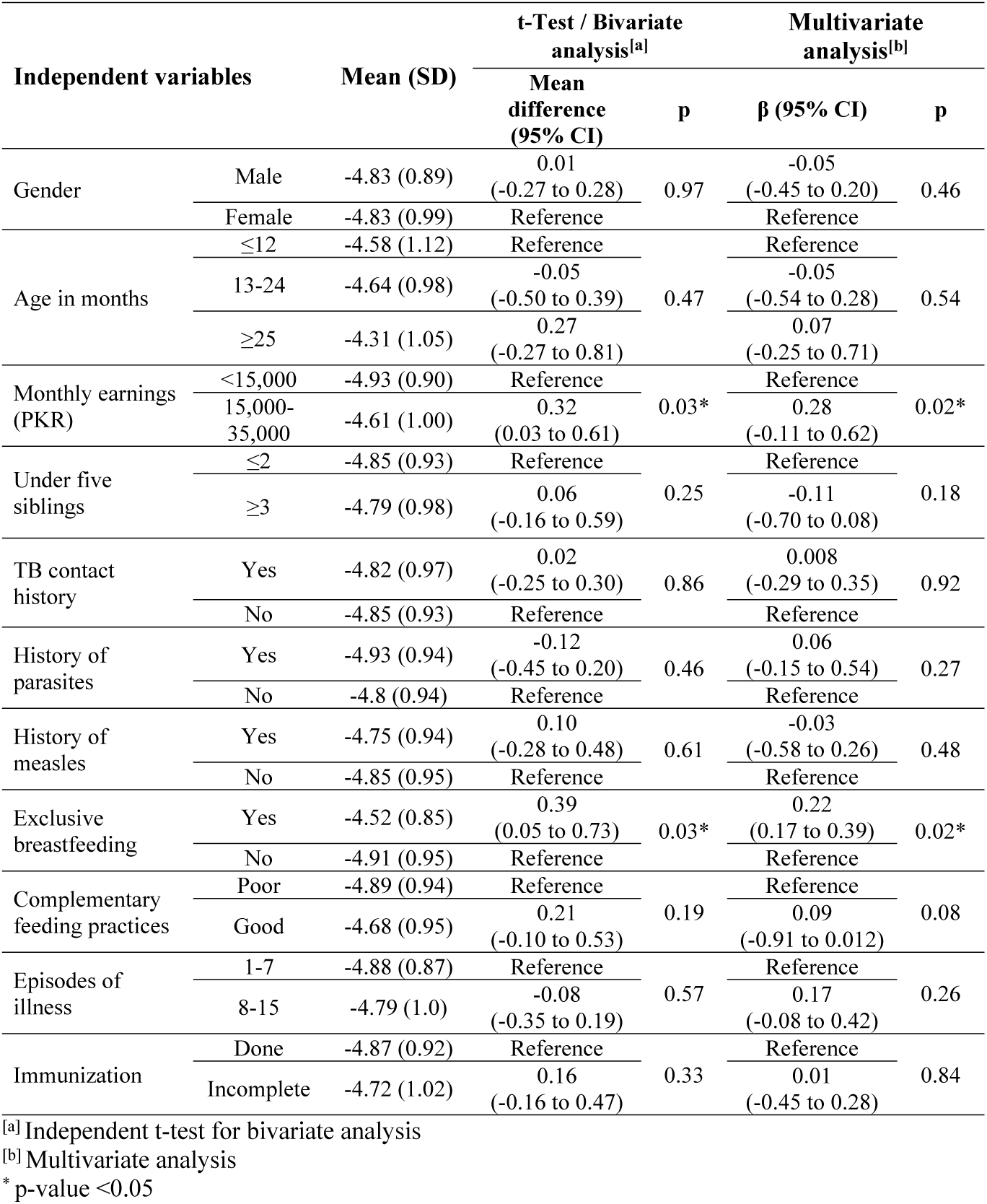
Determinants of undernutrition.

## Discussion

Malnutrition and undernutrition are detrimental for the future generation. Our study highlighted several determinants of wasting, stunting, and undernutrition. If addressed carefully, it could result in lower rates of children suffering from these conditions. Among others, the results show that children from low socio-economic status (monthly income less than 15,000 PKR) had a higher risk of wasting as compared to those children who belonged to relatively higher income (15,000 to 35,000 PKR monthly). Income level is an obvious factor that affects food intake. Families with low income are not able to feed their children properly due to limited resources. Some other studies also claim the effect of income on malnutrition [12,13].

In our study, the majority of the children were severely acute malnourished. In addition, chronic malnourishment owed to poor complementary feeding practices. Because weaning practices have a crucial role in the development of children and prevention of malnutrition, proper eating habits or good complementary feeding practices are key factors to improve the nutritional status of children. When children receive an adequate amount of food or a complementary diet, they have a positive trend towards healthy growth and development. Poor weaning practices negatively affect the nutritional status leading to malnutrition [2,9]. Complementary feeding along with breastfeeding up to 24 months of age provides an adequate amount of nutrients and also provides balanced nutrition that is essential not only for the development of children but also for the prevention of malnutrition [9].

Breastfeeding should start from birth and continue till the second birthday of the child along with complementary feeding that prevents acute and chronic malnutrition, as mother’s milk alone cannot fulfill the requirement of growing children [9,14,15]. Breastfeeding without complementary feeding after six months of age might be a risk factor for malnutrition due to the lack of essential nutrients that are necessary for the normal growth and development of children [9].

Improper and non-exclusive breastfeeding can lead to acute and chronic malnutrition. According to a study conducted in Pakistan, 48.4% of children were exclusively breastfed and 45.8% of mothers started early initiation of breastfeeding right after the birth of a baby [5]. Exclusive breastfeeding should start in the first hour after the birth, because mother’s milk has a balanced amount of nutrients necessary for the newborn [9,12]. Some infectious diseases have an impact on the degree of malnutrition as well as on the growth and development of children. Previous research has shown that children frequently having infections were more undernourished [16-18]. Children having a history of TB were more likely to be stunted and malnourished because of inflammation [16].

Children with three or more siblings are more likely to be stunted than those children who have one or two siblings. The point is that with more children in a household, lower amounts of food might be available per person. In this way, health and nutritional status may be compromised and children are more likely to be wasted or stunted [2,14,15]. Children born at birth intervals less than two years are more likely to develop acute or chronic malnutrition [11,14]. The higher the number of children under five in a family, the more scarce are the resources to properly feed all children [2,17,18]. A large household size also has been shown to negatively affect children’s health, nutritional status, and development [12,17].

Mother’s education has an important part in the healthy growth and development of infants and young children. In the current, the study majority of mothers were uneducated and their children showed more nutritional problems. An educated mother may be better aware of the nutritional needs of her child and, thereby, may prevent stunting and wasting [19]. There are some limitations to this study due to its cross-sectional design. Anthropometric measures have been conducted by qualified personell which promote data quality. By observing children’s nutritional state with greater insight, the dynamic nature of child’s growth and development could be better understood.

## Conclusion

Children under the age of five years are prone to malnutrition as well as undernutrition. Almost all developing countries are suffering from the public health issues of malnutrition as well as undernutrition among children. The findings of the current study indicate that wasting and stunting along with undernutrition are associated with exclusive breastfeeding and complementary feeding practices, household size, family earning, and history of infections. Concerted efforts will be needed from all sectors of society to tackle the problem of child malnutrition in Pakistan. Alertness regarding a child’s dietary needs and other factors leading to child growth must be incorporated in the child nutrition program.

## Data Availability

The study is based on clinical data from Community Management of Acute Malnutrition (CMAM) centers which cannot be published. However, the data is available from the corresponding author upon reasonable request.

## List of abbreviations

CI: Confidence interval
CMAM: Community Management of Acute Malnutrition
EPI: Expanded Program of Immunization
PKR: Pakistani rupee
SD: Standard deviation
TB: Tuberculosis
WHO: World Health Organization

## DECLARATIONS

### Ethical approval and consent to participate

The study was conducted after the approval of the Ethical Review and Advanced Study Research Board of the University of Punjab, Pakistan (ref-9/2352-ACAD).. Written consent was obtained from parents after introducing them to the purpose of the study.

### Consent for publication

Not applicable

### Availability of data and materials

Data is available from corresponding author upon reasonable request.

### Author contributions

Conceptualization: J.S. and R.Z.; Data curation: J.S.; Formal analysis: J.S.; Writing – original draft: J.S.; Writing – reviewing & editing: R.Z., R.M.A., M.S.B., F.M., G.M.J.B. and F.F.; supervision: G.M.J.B. and F.F. All authors read and approved the final version of the manuscript.

## Acknowledgments

Not applicable.

## References

1. World Health Organization. Malnutrition Fact Sheet. Available online: https://www.who.int/news-room/fact-sheets/detail/malnutrition; 2021 [accessed 22 October 2021].

2. UNICEF, WHO, World Bank Group. Levels and trends in child malnutrition. UNICEF / WHO / World Bank Group Joint Child Malnutrition Estimates. Key findings of the 2020 edition. Geneva: World Health Organization; 2020.

3. Hossain FB, Shawon MS, Al-Abid MS, Mahmood S, Adhikary G, Bulbul MM. Double burden of malnutrition in children aged 24 to 59 months by socioeconomic status in five South Asian countries: evidence from demographic and health surveys. BMJ Open. 2020;10(3):e032866.

4. Waber DP, Bryce CP, Girard JM, Zichlin M, Fitzmaurice GM, Galler JR. Impaired IQ and academic skills in adults who experienced moderate to severe infantile malnutrition: a 40-year study. Nutritional Neuroscience. 2014;17(2):58–64.

5. UNICEF. National Nutrition Survey 2018 – Key Findings Report. Available online: https://www.unicef.org/pakistan/reports/national-nutrition-survey-2018-key-findings-report; 2019 [accessed 22 October 2021].

6. Demographic and Health Surveys. Calverton: MEASURE DHS. Pakistan; 2013.

7. World Health Organization. Guideline: updates on the management of severe acute malnutrition in infants and children. Geneva: World Health Organisation; 2013.

8. World Health Organization. WHO child growth standards and the identification of severe acute malnutrition in infants and children: a joint statement by the World Health Organization and the United Nations Children’s Fund. Geneva: orld Health Organization; 2009.

9. World Health Organization. Infant and Young Child Feeding. Available online: https://www.who.int/news-room/fact-sheets/detail/infant-and-young-child-feeding; 2021 [accessed 22 October 2021].

10. Waheed M. WHO EMRO | Expanded Programme on Immunization | Programmes | Pakistan. Geneva: World Health Organization; 2020.

11. Selina L, Pamela D, Richard H, Lancet Early Childhood Development Series Steering Committee. A good start will ensure a sustainable future for all. Lancet. 2017;389:8–9.

12. Nambuusi BB, Ssempiira J, Makumbi FE, Kasasa S, Vounatsou P. The effects and contribution of childhood diseases on the geographical distribution of all-cause under-five mortality in Uganda. Parasite Epidemiology and Control. 2019;5:e00089.

13. Nott J. Malnutrition in a modernizing economy: The changing aetiology and epidemiology of malnutrition in an African Kingdom, Buganda c. 1940–73. Medical History. 2016;60(2):229–249.

14. Kahssay M, Woldu E, Gebre A, Reddy S. Determinants of stunting among children aged 6 to 59 months in pastoral community, Afar region, North East Ethiopia: unmatched case control study. BMC Nutrition. 2020;6:9.

15. Eunice MJ, Cheah WL, Lee PY. Factors Influencing Malnutrition among Young Children in a Rural Community of Sarawak. Malaysian Journal of Nutrition. 2014;20(2):145–164.

16. Wagstaff A, Bustreo F, Bryce J, Claeson M. Child health: reaching the poor. American Journal of Public Health. 2004;94(5):726–736.

17. Chattopadhyay N, Saumitra M. Developmental Outcome in Children with Malnutrition. Journal of Nepal Paediatric Society. 2016;36(2):170–177.

18. Gupta N, Kabra M. Approach to the diagnosis of developmental delay - the changing scenario. Indian Journal Medical Research. 2014;139:4–6.

19. Mason F, Rawe K, Wright S. Superfood for babies: how overcoming barriers to breastfeeding will save children’s lives. London: Save the Children; 2013.

